# Structural disconnectome mapping of cognitive function in post-stroke patients

**DOI:** 10.1101/2021.06.25.21259526

**Authors:** Knut K. Kolskår, Kristine M. Ulrichsen, Genevieve Richard, Erlend S. Dørum, Michel Thiebaut de Schotten, Jaroslav Rokicki, Jennifer Monereo-Sánchez, Andreas Engvig, Hege Ihle Hansen, Jan Egil Nordvik, Lars T. Westlye, Dag Alnæs

## Abstract

Sequalae following stroke represents a significant challenge in current rehabilitation. The location and size of focal lesions are only moderately predictive of the diverse cognitive outcome after stroke. One explanation building on recent work on brain networks proposes that the cognitive consequences of focal lesions are caused by damages to anatomically distributed brain networks supporting cognition rather than specific lesion locations. To investigate the association between post-stroke structural disconnectivity and cognitive performance, we estimated individual level whole-brain disconnectivity probability maps based on lesion maps from 102 stroke patients using normative data from healthy controls. Cognitive performance was assessed in the whole sample using Montreal Cognitive Assessment, and a more comprehensive computerized test protocol was performed on a subset (n=82). Multivariate analysis using Partial Least Squares on the disconnectome maps revealed that higher disconnectivity in right insular and frontal operculum, superior temporal gyrus and putamen was associated with poorer MoCA performance, indicating that lesions in regions connected with these brain regions are more likely to cause cognitive impairment. Furthermore, our results indicated that disconnectivity within these clusters was associated with poorer performance across multiple cognitive domains. These findings demonstrate that the extent and distribution of structural disconnectivity following stroke are sensitive to cognitive deficits and may provide important clinical information predicting post stroke cognitive sequalae.

## Introduction

The magnitude and characteristics of cognitive impairments following stroke show substantial individual differences across patients. While some patients show considerable deficits and subsequent increased risk of neurodegeneration and dementia, others show no observable impairments. This heterogeneity in cognitive deficits after stroke arise not only from differences in the localization and extent of focal lesions, but also from potential secondary cascade effects in terms of altered brain connectivity (Rehme & Grefkes, 2013), and alterations of the hierarchical brain network structure (Stam, 2014). Currently, common predictors of cognitive deficits and recovery include anatomical location, lesion severity, vascular risk factors, chronic brain pathology and pre-stroke cognitive impairment (Bentley et al., 2014; Macciocchi, Diamond, Alves, & Mertz, 1998; Munsch et al., 2016; Pendlebury, 2009). However, recent studies indicate added predictive value of connectivity-based measures, which capture perturbations of brain network connections or dynamics beyond focal lesion site (Ktena Sofia et al., 2019; Lopes et al., 2021b).

In the last decade, a large body of literature has characterized the brain as a complex network consisting of nodes and their connections, collectively termed the brain connectome. Building on functional imaging, a coarse parcellation of networks separate the brain in the cingulo-opercular, frontoparietal, ventral attention and default mode networks, aiding cognitive control. The cingulo-opercular network is associated with the ability to maintain focus, the ability to sustain top-down cognitive control across cognitive tasks (Cai et al., 2016; Hilger, Ekman, Fiebach, & Basten, 2017; Uddin, Yeo, & Spreng, 2019; Wilk, Ezekiel, & Morton, 2012), whereas the frontoparietal network is assumed to guide moment to moment-attentional control (Fassbender et al., 2006; Majerus, Péters, Bouffier, Cowan, & Phillips, 2018). The ventral attention network serves reorientation to relevant stimuli outside the scope of current attention (Maurizio Corbetta & Gordon L Shulman, 2011; Vossel, Geng, & Fink, 2014), while default mode network is commonly associated with self-referential internal activity, and is commonly downregulated during task engagement (McKiernan, Kaufman, Kucera-Thompson, & Binder, 2003; Vatansever, Manktelow, Sahakian, Menon, & Stamatakis, 2018). A central property of network nodes is how densely they are connected. Highly connected nodes, or hubs, are critical for efficient information flow between brain regions and are thought to play a crucial role in pivoting neural activity across distal brain regions, allowing for the integration of information required to support cognitive operations (Cole et al., 2013). Thus, focal damage to any part of a brain network might cause disruptions of distal but intact brain regions. Indeed, even small lesions in a densely connected hub may cause connectome-wide perturbations (Aben et al., 2019). Recent studies have therefore moved beyond traditional lesion-symptom mapping, to include measures of network dysfunction to explain and predict stroke sequelae (Lim & Kang, 2015; Ulrichsen et al., 2020). Recent findings using the lesion-network mapping approach suggest that patients with overlapping symptoms have lesions in regions that are functionally connected and that lesions to brain network hubs or their connecting white matter pathways are associated with more symptoms (Fox, 2018). This network-based concept has also been shown to apply to a wide range of brain disorders such as Alzheimer disease, schizophrenia and multiple sclerosis (Crossley et al., 2014; Stam, 2014; van den Heuvel & Sporns, 2019).

Whereas association between functional connectivity following stoke has been investigated in recent years (Klingbeil, Wawrzyniak, Stockert, & Saur, 2019; Lopes et al., 2021a; Ptak et al., 2020), less is known regarding structural connectivity. The brain is connected by an intricate web of white matter (WM) pathways, consisting of bundles of myelinated axons responsible for conveying signals between brain regions, supporting functional networks. While measuring axonal disconnections in the living human brain has remained a challenging task, a recent implementation of diffusion tensor imaging (DTI) based tractography for assessing full-brain connection probabilities has enabled opportunities for detailed estimation of the connections of one or several lesions in individual patients (Foulon et al., 2018). Based on normative data from healthy controls, voxel-wise disconnection probability maps for a particular patient can be derived based on a lesion map. The extent and distribution of these disconnectivity maps have been shown to correlate with a surrogate biomarker of neuronal damage in patients with MS (Rise et al., 2021) and shown promise in predicting deficits following stroke (Salvalaggio, De Filippo De Grazia, Zorzi, Thiebaut de Schotten, & Corbetta, 2020).

To test for associations between brain disconnection and cognitive performance, we used Partial Least Squares analysis (PLS) to map common variance between voxel-wise structural disconnection probability maps in 102 stroke survivors and their performance on the Montreal Cognitive assessment (MoCA; Nasreddine et al., 2005). PLS (Krishnan, Williams, McIntosh, & Abdi, 2011) is well-suited for investigating multivariate associations between neuroimaging features and non-gaussian behavioral and clinical measures commonly obtained from stroke patients (Blackburn, Bafadhel, Randall, & Harkness, 2013). Although PLS has been suggested to display less anatomical specificity, it has shown to produce higher stability in results (Ivanova, Herron, Dronkers, & Baldo, 2021). Importantly, MoCA have been shown to reliably identify cognitive impairment in clinical samples (Bernstein, Lacritz, Barlow, Weiner, & DeFina, 2011), across latent variables related to executive function, language, memory visuospatial skills, working memory, as well as orientation (Freitas, Simões, Marôco, Alves, & Santana, 2012). Based on the literature reviewed above, we hypothesized that patients with higher levels of stroke-induced brain disconnectivity would show poorer cognitive performance, and further anticipated that this association could not simply be explained by the size and location of the lesion itself. We further hypothesized that derived scores from the PLS would correlate with a broader cognitive battery derived for stroke patients (Willer, Pedersen, Forchhammer, & Christensen, 2016). Since previous studies and existing models of the distributed nature of the neuroanatomical basis of cognitive functions are sparse, we remained agnostic about the anatomical distribution of the associations with the disconnectome maps and performed an unbiased full brain analysis with appropriate corrections for multiple comparisons.

## Methods

The present cross-sectional study included participants previously described in detail (Dørum et al., 2020; Kolskår et al., 2020; Richard et al., 2020; Ulrichsen et al., 2020). Briefly, the sample comprised 102 stroke survivors from sub-acute (>24h post stroke and in clinical stable condition) to chronic stage. Inclusion criteria were radiologically documented ischemic or hemorrhagic stroke, and exclusion criteria were psychiatric conditions (bipolar disorder or schizophrenia), other neurological conditions including known cognitive impairment pre-stroke, substance abuse, and contraindications for MRI compatibility. All participants gave their written consent before participating, and the study was approved by the Regional Committee for Medical and Health Research Ethics South-East Norway (2014/694 and 2015/1282).

Table 1 displays sample demographics as well as time between stroke and MRI and cognitive assessment.

**Table 1.** Summary of clinical and demographic variables. NIHSS: National Institute of Stroke Scale. CapPad: Cognitive Assessment at Bedside for iPad. Acute: Less than seven days since stroke. Subacute: between 7 and 180 days since stroke. Chronic: more than 180 days since stroke. * Based on radiological description. Number of lesions and lesion size is based on MNI-normalized lesion masks.

|  | Mean | sd | min | max |
| --- | --- | --- | --- | --- |
| Age | 66.3 | 12.3 | 24 | 87 |
| Sex | 74.5% males |  |  |  |
| Education (self-report, years) | 14.6 | 3.3 | 7 | 30 |
| Interval between stroke onset and MRI / MoCA assessment (days) | 515 | 444 | 1 | 1399 |
| Lesion size (2mm resolution) | 7693 | 24723 | 5 | 30031 |
| Number of lesions | 3 | 2 | 1 | 12 |
| Interval between stroke onset and CabPad- assessment (days) | 634 | 379 | 89 | 1428 |
| NIHSS (score) | 1.02 | 1.35 | 0 | 7 |
| Hemispheric distribution Left:39 / Right:41 / Bilateral:19 / Subcortical: 2 |  |  |  |  |
| Time since stroke: Acute: 20 / Subacute: 25 / Chronic:57 |  |  |  |  |

### Cognitive assessment

Patients were assessed with MoCA (Nasreddine et al., 2005) at time of MRI scanning. At follow-up (Table 1), a subsample (n=82) were also assessed using CabPad (Willer et al., 2016), a computerized test battery assessing a range of functions, including motor speed (finger tapping) for dominant and non-dominant hand, verbal fluency (phonetic and semantic word generation), attention span (symbol sequencing), working memory (reversed symbol sequencing), a spatial Stroop / flanker task, spatial short-term memory and psychomotor speed (symbol-digit coding task).

### MRI acquisition

Patients were scanned at Oslo University Hospital on a 3T GE 750 Discovery MRI scanner with a 32-channel head coil. T2-FLAIR images were acquired with the following parameters: TR: 8000 ms; TE: 127 ms, TI: 2240 ms; flip angle (FA): 90°; voxel size: 1×1×1 mm. T1-weighted scans were collected using a 3D IR-prepared FSPGR (BRAVO) sequence (TR: 8.16 ms; TE: 3.18 ms; TI: 450 ms; FA: 12°; voxel size: 1×1×1 mm; FOV: 256 × 256, 188 sagittal slices) for co-registration.

### Lesion delineation

Lesion delineation was initially guided on each participant’s FLAIR image, using the semi-automated Clusterize-Toolbox, implemented in MATLAB (de Haan, Clas, Juenger, Wilke, & Karnath, 2015), before finalized by manual demarcation. The demarcation was guided by radiological descriptions, and evaluated by a medical doctor before finalization. Normalization parameters were estimated by linear registration of the structural T1-image to the MNI152-template and applied to the lesion masks using Flirt (Jenkinson & Smith, 2001). Figure 1 displays overlap in lesion location across the sample.

**Figure 1.**
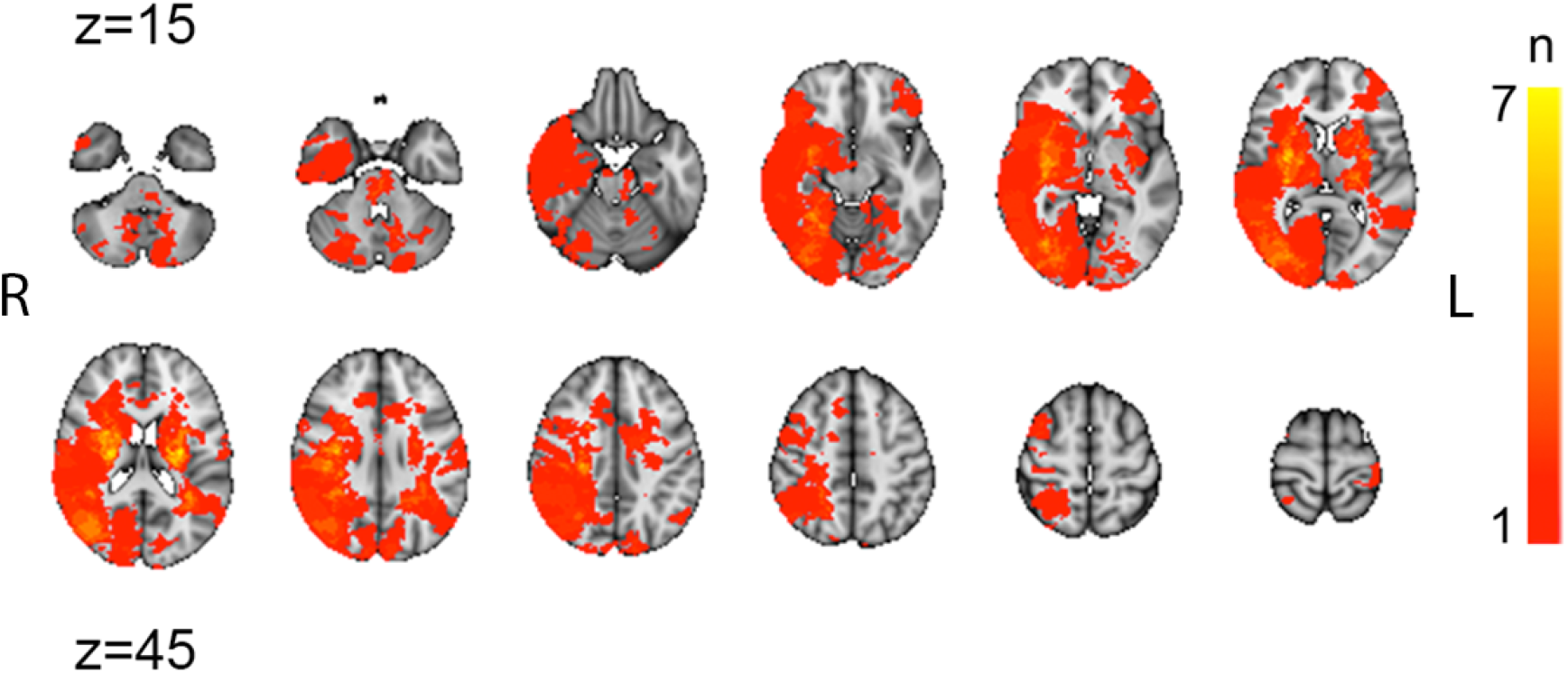
Lesion overlap across 102 stroke patients. 12 transversal slices, with 5 mm thickness. Color scale indicates number of participants overlapping. Z coordinates denotes transversal slices in the MNI152-coordinate system.

### Group comparisons

To asess goup differences in MoCA-score between the patients in acute, subacute and chronic phase, a linear model was estimated with MoCA-score as dependent variable, and group as independent variable. To assess impact of lesion load, a linear model was estimated with MoCA-score as dependent variable and lesion size and number of lesions as independent variables.

### Estimation of structural disconnectome maps

To estimate the extent of the structural disconnection for each patient, we employed an automated tractography-based procedure (Foulon et al., 2018). Briefly, full-brain tractography data of 170 healthy controls from the 7T Human Connectome Project was used as a normative training set to identify fibers passing through each lesion (Thiebaut de Schotten, Foulon, & Nachev, 2020). Using affine and diffeomorphic deformations (Avants et al., 2011; Klein et al., 2009), individual patient’s lesion maps were registered to each control’s native space and used as seeds for the probabilistic tractography in Trackvis (Wang, Benner, Sorensen, & Wedeen, 2007). The resulting tractograms were transformed to visitation maps, binarized, and registered to MNI152 space before a percentage overlap map was produced by summarizing each point in the normalized healthy subject visitation maps. The resulting disconnectome maps are whole-brain voxel-wise probability maps indicating for each patient and for each voxel in the brain the probability that the voxels were disconnected. Next, these individual-level disconnection maps were included in group-level analysis.

### Statistical analysis

To assess associations between MoCA scores and disconnectome maps, we used non-rotated task-based PLS using PLS Application (Krishnan, Williams Lj Fau - McIntosh, McIntosh Ar Fau - Abdi, & Abdi, 2010) for MATLAB (MathWorks, 2018), entering female, age and MoCA score as behavioral variables, with contrasts for mean effects for each (1 0 0; 0 1 0; 0 0 1). We performed permutations (n=1000) to assess the significance of the estimated latent variables, while precision was estimated using bootstrapping (n=1000) and used to calculate pseudo-z or bootstrap ratio brain maps (McIntosh & Lobaugh, 2004).

To assess associations with cognitive performance, disconnectivity within each significant PLS cluster associated with MoCA was extracted and correlated with CabPad performance.

## Results

Figure 2 displays patient distribution of MoCA-scores. 35% of the patients fulfilled criteria for mild cognitive impairment, based on a suggested cutoff at 26 (Nasreddine et al., 2005).

**Figure 2.**
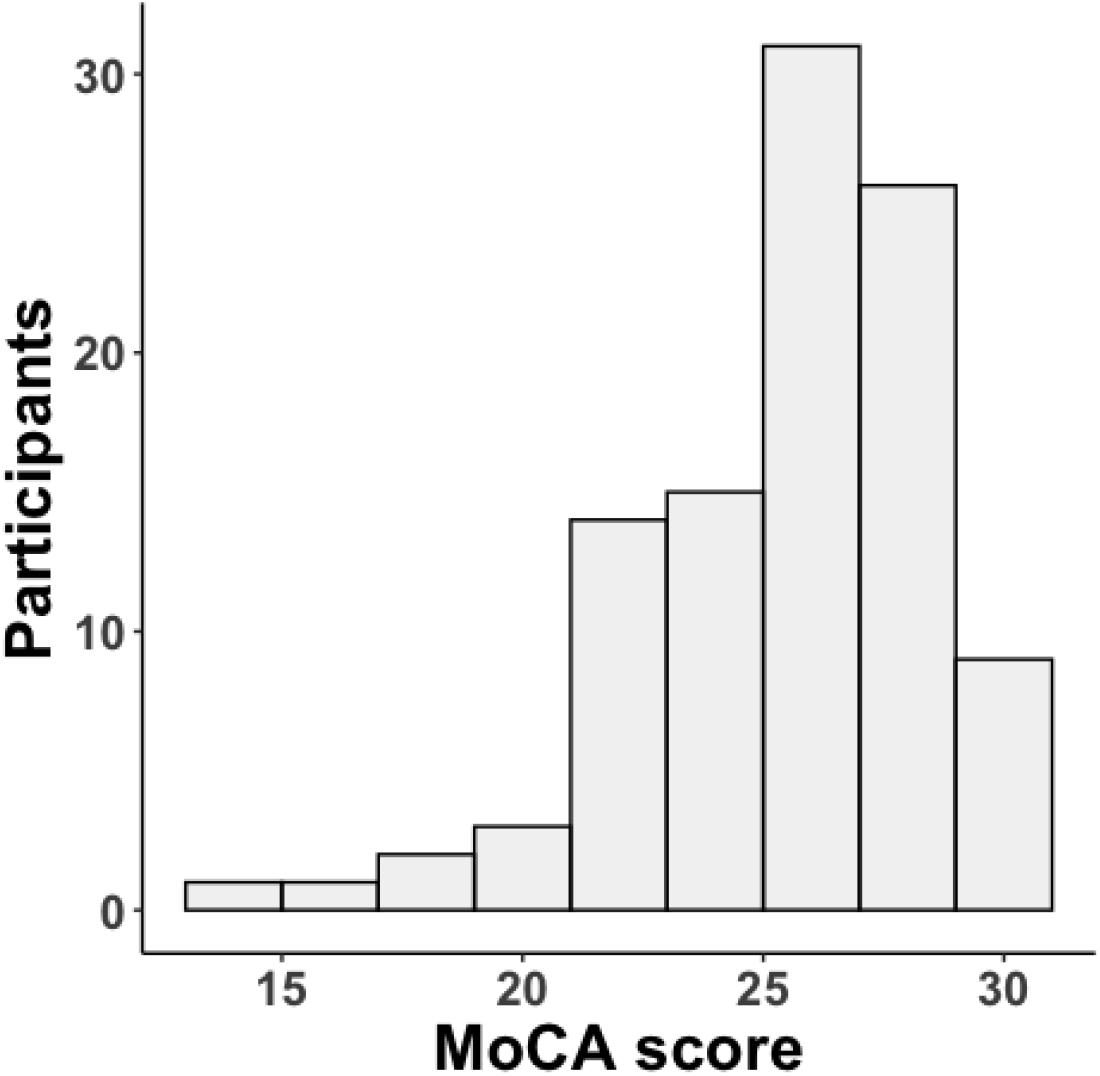
Histogram depicting the distribution of MoCA scores across the sample.

Investigation of association between group and MoCA performance revealed no significant relationship (F=1.8, p=0.17, Figure 3). All participants were therefore pooled for further analysis. Investigation of association between MoCA score and lesion load identified a significant association between total lesion volume and MoCA score (t=-2.96, p=0.004), but not with number of lesions (t=-1.502, p=0.13). However, after removal of one participant with an extreme lesion size, the association between MoCA and lesion volume did not remain significant (t=-1.7, p=0.09).

**Figure 3.**
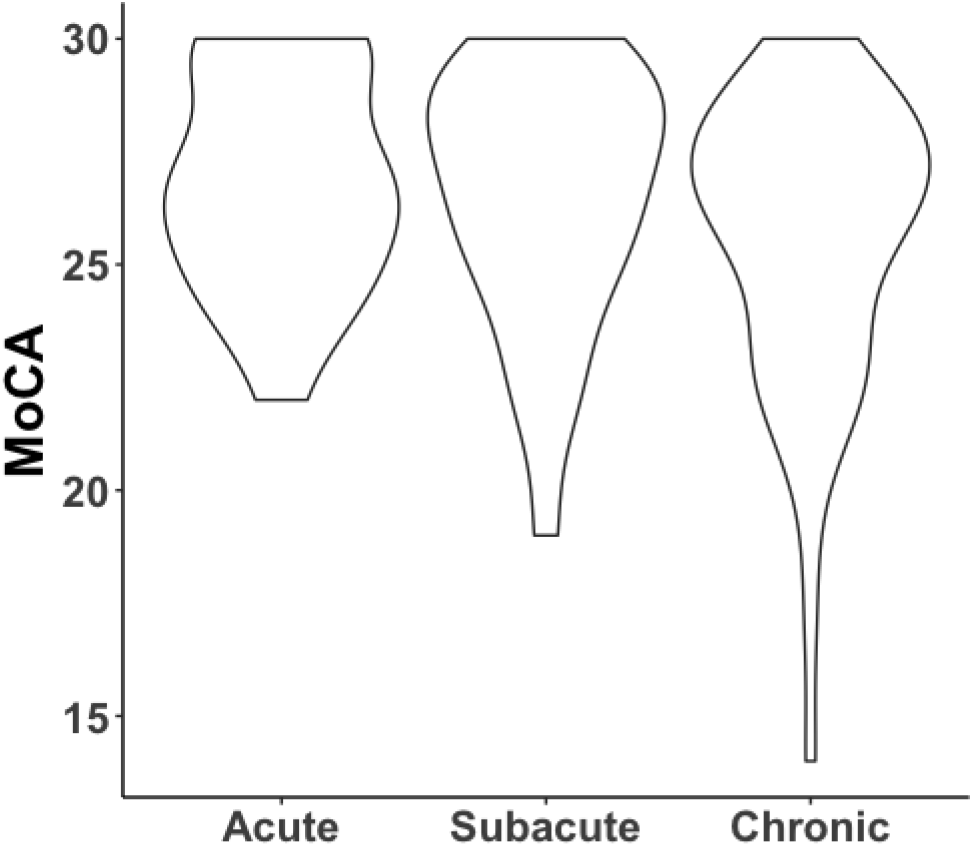
Distribution of MoCA scores by patient group.

Figure 4 and Table 2 summarize the results from the PLS. Briefly, permutation testing revealed three significant clusters (pseudo-z > 3) showing common variance between MoCA performance and structural disconnectivity, including 1) right frontal operculum/insular cortex, 2) right superior temporal gyrus, and 3) the right putamen. Of note, association between lesion size and individual PLS brain scores remained highly similar after removal of an individual with an extreme lesion size (included: t=-9.1, p= 7.6e-15, excluded: t=-7.4 p=5.13e-11), indicating that our main results within the PLS was not strongly driven by the outlier.

**Figure 4.**
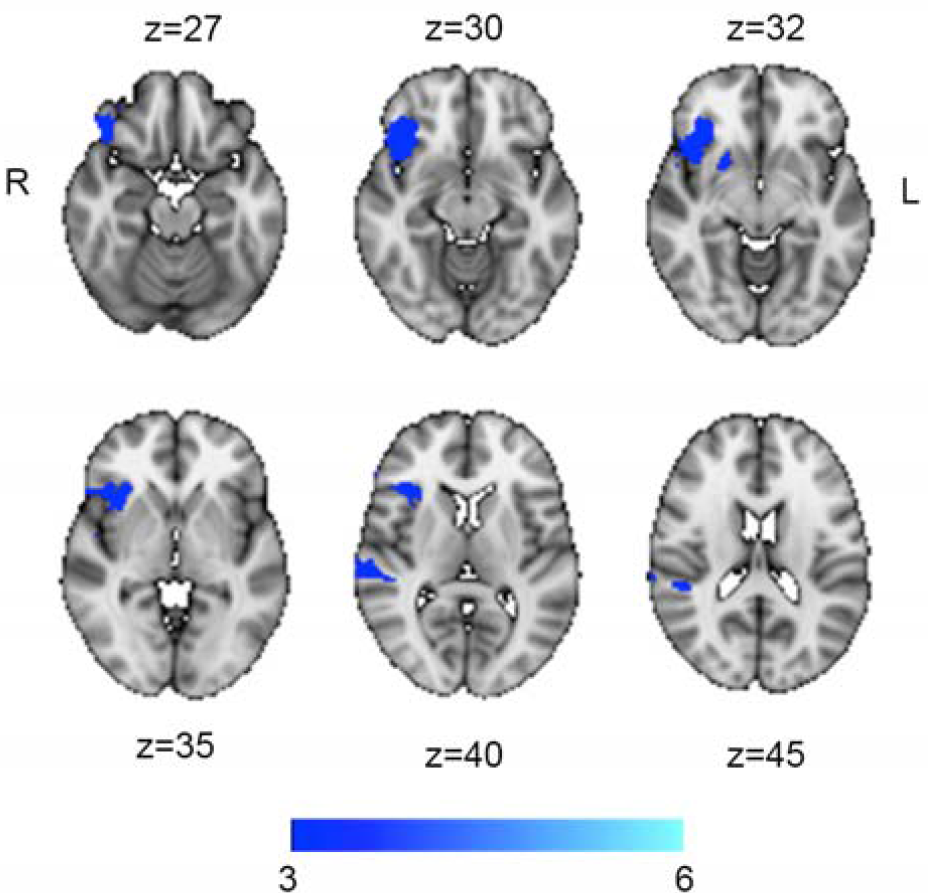
Voxel-wise disconnectome bootstrap ratio maps for the MoCA association, thresholded at pseudo-z >3.

**Table 2.** Disconnectome clusters significantly associated with MoCA performance, identified by PLS analysis.

| Cluster-# | Location | Size $\text{mm}^3$ | Max pseudo-Z | X, Y, Z (vox) |
| --- | --- | --- | --- | --- |
| 1 | Right frontal operculum/insula | 1063 | 4.09 | [25, 75, 33] |
| 2 | Right superior temporal gyrus, posterior division | 273 | 3.65 | [15, 48, 41] |
| 3 | Right putamen | 37 | 3.61 | [34, 68 32] |

Figure 5 illustrates correlations between disconnectivity within each of the significant clusters identified by MoCa-PLS and CabPad performance. Disconnectivity was negatively associated with performance on all tests, with strongest correlations for semantic and phonetic fluency.

**Figure 5.**
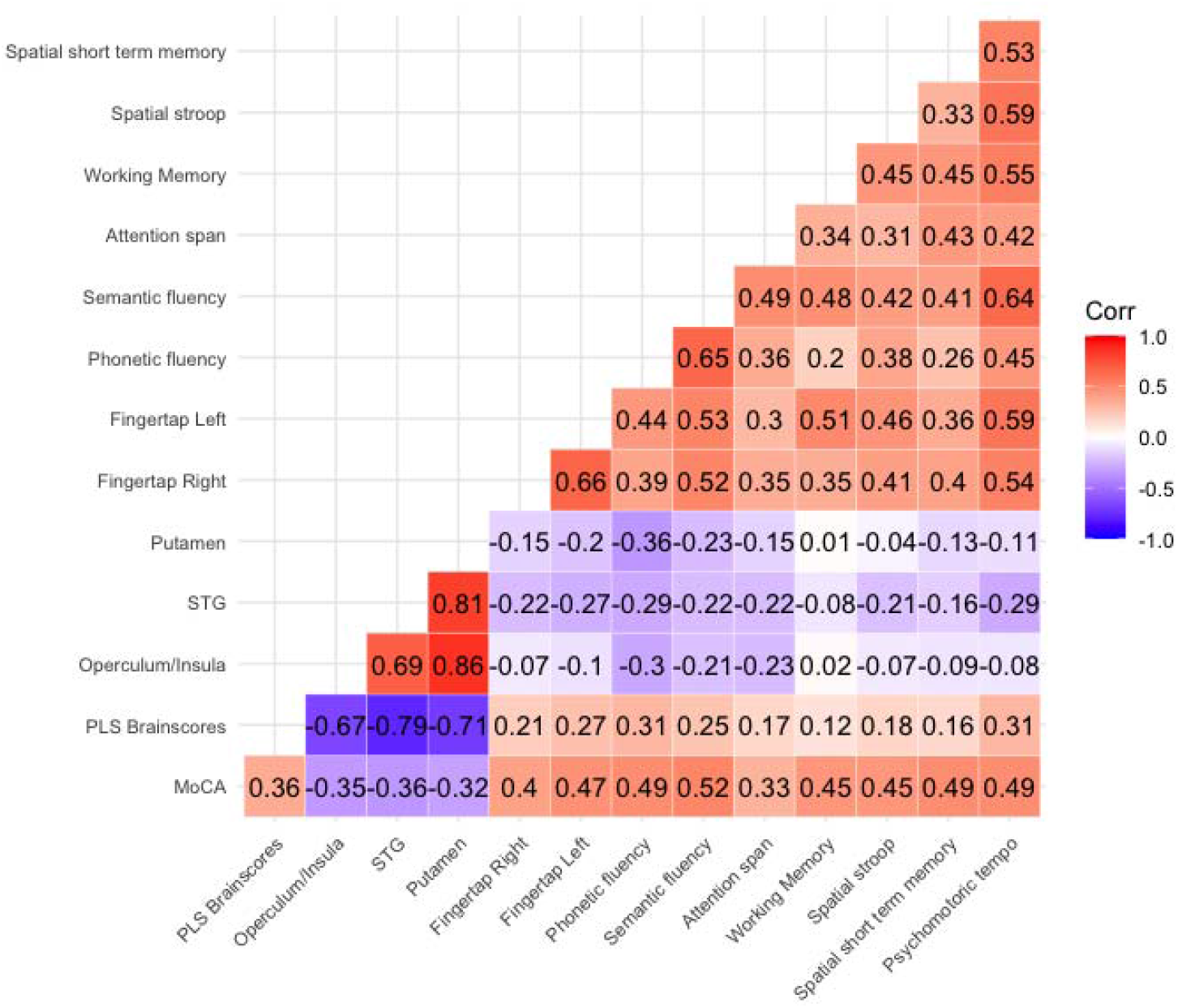
Correlation between MoCA, PLS-weights, disconnectivity within the significant clusters, and CabPad performance.

## Discussion

In the current study, we investigated associations between individual structural disconnectome maps and post-stroke cognitive performance as measured with MoCA in 102 stroke survivors. We also performed follow-up investigation assessing cognitive associations with a broader cognitive battery on a subsample. Our analysis revealed that structural disconnections implicating the right insula and frontal operculum, along with right superior temporal gyrus and right putamen were associated with poorer general cognitive performance as measured with MoCA. In a subsample comprising 82 of the patients, we demonstrated that the association generalized to a range of cognitive domains, including word generation, attention span, and speed. Together, our results support the relevance of altered structural connectivity on cognitive impairments following stroke.

Stroke frequently cause cognitive impairment, and investigations of the underlying network dynamics may add to our current understanding and prognostics. Structural disconnections caused by stroke have been shown to influence functional connectivity both directly and indirectly (Griffis, Metcalf, Corbetta, & Shulman, 2019, 2020). This suggests a mechanism of cognitive sequelae in which altered functional connectivity due to structural disruptions cause aberrant function in distal nodes, where lesions perturbating any part of a network supporting cognitive functions may cause cognitive and behavioral impairment (Alstott, Breakspear, Hagmann, Cammoun, & Sporns, 2009) (Chen et al., 2019; Lim & Kang, 2015).

A challenge in current healthcare is assessment of cognitive function with a sensitivity and specificity able to detect and differentiate cognitive difficulties (Dong et al., 2010). MoCA (Nasreddine et al., 2005) has been suggested as a feasible screening tool for detecting cognitive impairment following stroke (Horstmann, Rizos T Fau - Rauch, Rauch G Fau - Arden, Arden C Fau - Veltkamp, & Veltkamp, 2014; Julayanont & Nasreddine, 2017; Munthe-Kaas et al., 2021), where key advantages are short administration time and multiple cognitive domains of assessment (Burton & Tyson, 2015; Stolwyk Renerus, O’Neill Megan, McKay Adam, & Wong Dana, 2014). Importantly, MoCA is sensitive to specific cognitive domains such as attention, executive and visuospatial function when utilized in lesion-mapping studies (Shi et al., 2018; Zhao et al., 2017). Of note, post stroke MoCA score has been demonstrated to differentiate cerebral blood flow in anterior cingulate and prefrontal cortex (Nakaoku et al., 2018). Anterior cingulate is strongly connected to insula and frontal operculum both structurally (Ghaziri et al., 2017) and functionally (Horn et al., 2010), and is suggested to be among the core nodes of the cingulo-opercular network. Importantly, these network nodes are suggested to mediate regulation and differentiation between the frontoparietal, ventral attention and default mode networks (Goulden et al., 2014; Menon & Uddin, 2010). The frontoparietal and default mode network are furthermore viewed as anticorrelated networks reflecting cognitive state, where degree of this differentiation have been shown to predict cognitive function in stroke survivors at the group (Geranmayeh, Leech, & Wise, 2016) and individual level (Lorenz et al., 2021). Indeed, lesions causing altered connectivity in cingulo-opercular nodes have been linked to cognitive impairment across multiple domains (Siegel et al., 2016; Warren et al., 2014), including general impairments as measured using MoCA (Vicentini et al., 2021).

Our analysis revealed significant associations between MoCA and disconnectivity in the right superior temporal gyrus (STG) and a small cluster in the putamen. STG is commonly associated with language production and interpretation (Brugge, Volkov, Garell, Reale, & Howard, 2003), and has also been found to be activated in in conjunction with insula during evaluation of responses in decision-making tasks (Megías, Cándido, Maldonado, & Catena, 2018; Paulus, Feinstein, Leland, & Simmons, 2005), where information needs to be integrated and evaluated over longer periods. STG has also been found to coactivate with visual attention, frontoparietal and cingulo-opercular nodes during reorientation of visual attention (Vossel et al., 2014), suggesting a more general role in attentional processing.

Putamen is involved in various aspects of motor functioning and learning, including language functions and reward signaling. The putamen is connected to thalamic and motor cortices (Jung et al., 2014; Leh, Ptito, Chakravarty, & Strafella, 2007), and is strongly connected to the frontoparietal network where the integrity of the connecting pathway is correlated with executive functioning in healthy adults (Bennett & Madden, 2014; Ystad et al., 2011). These results together suggests that post-stroke cognitive dysfunction may partly arise from lack of differentiation between large-scale brain networks. Our results align with this understanding, as structural disconnectivity of the insula and operculum was associated with impaired MoCA performance, potentially through disrupted network regulation.

Expanding our results, disconnectivity within the MoCA-associated clusters correlated with performance across several cognitive domains as measured by CabPad. While the apparent lack of cognitive specificity within and across clusters may reflect that the disconnectome approach captures a more over-arching impairment, strongest correlations were seen with phonetic and semantic generation and visual attention span. Phonetic and semantic fluency reflect executive functioning beyond pure language generation (Delis, Kramer, Kaplan, & Holdnack, 2004), and greater functional connectivity within the cingulo-opercular and ventral attention network has been associated with reading fluency.

Our results revealed significant associations within the right hemisphere only. A recent investigation of structural connectivity in patients with ischemic leukoaraiosis found predominantly right side altered graph metrics when compared to healthy controls, and reported associations with cognitive performance (Lu et al., 2021). Comparably, right side functional abnormalities in cingulo-opercular nodes during resting state have been associated with impaired MoCA performance following a transient ischemic attack (Guo et al., 2014), and right side dominance in bilateral insula activation is robustly found during task engagement in healthy controls (Rottschy et al., 2012). Further, right hemisphere is to a larger degree linked to visual attention, reorientation, and bottom-up processing, and damage to right side pathways associated with impaired spatial attention, target detection and vigilance (Maurizio Corbetta & Gordon L. Shulman, 2011). Albeit speculative, our results indicate a larger right-side vulnerability for abrupted structural connectivity augmenting post stroke cognitive difficulties.

The current study has limitations. First, our cross-sectional design does not allow for integrating premorbid cognitive function and chronic brain pathology, which is highly relevant as an outcome predictor (Sagnier & Sibon, 2019). Further, the current sample represents a heterogeneous group regarding stroke severity, we lack measures on vascular risk and the sample size is moderate, factors that are relevant for the generalizability of the reported findings to the stroke population in general (Marek et al., 2020). Of note, our data did not allow for differentiation between stroke reported at hospital admittance and potential older strokes reported in the radiological description. We can therefore not rule out the possibility that the associations are partly driven by previous strokes in some of the patients. Further, jointly estimating all voxels in a single PLS model increases sensitivity, however, comes with a trade-off in spatial specificity as we can only make statements about significance for the spatial pattern as a whole. PLS has shown comparable performance compared to similar multivariate lesion symptom mapping approaches, it has shown to display less anatomical specificity, warranting caution when interpreting spatially sparse clusters (Ivanova et al., 2021). Still, the reliability of each voxel’s contribution to the observed pattern was assessed through bootstrapping. While the follow-up association between brain scores and the broader CabPad-battery may indicate whether the MoCa-driven PLS-decomposition is driven by general or more specific cognitive domains, it cannot capture novel associations not initially captured by the MoCA Although MoCA has been shown to display adequate sensitivity and specificity to detect mild cognitive impairment, it does not offer a comprehensive assessment. Indeed, lack of specificity and variability in difficulty increases the likelihood to overlook minor cognitive difficulties. Indeed, MoCA only provide a crude proxy for cognitive impairment, and does not allow for drawing interference on broad cognitive abilities. Of note, premorbid cognitive function is associated with post stroke outcome. In the current data collection we did not obtain measures allowing for estimation of premorbid function” In conclusion, our study supports the relevance of investigating disrupted structural connectivity following stroke. Although our study does not allow differentiation between proximal and distal effects due to sample size and lesion homogeneity, our results highlight the relevance of altered structural connectivity when investigating cognitive sequalae after stroke. In line with previous studies, our results indicate lesions affecting the insula and the frontal operculum are associated with cognitive impairment and support the inclusion of measures of structural disconnection when evaluating cognitive and functional sequelae in stroke patients.

## Data Availability

Open science and data availability statement:
Relevant scripts and scrambled and anonymized data needed to reconstruct the reported analyses and results are available on OSF (link below). While the sensitive nature of the data and our current approvals do not allow for public sharing of real data, anonymized data may be available upon request to the corresponding author following appropriate data transfer agreements.
OSF-repository: https://osf.io/juwhv/?view_only=1096394b4b2b4a51abceea5834d29969.

https://osf.io/juwhv/?view_only=1096394b4b2b4a51abceea5834d29969

